# AI-Enhanced Memory List Assessment System: Multi-Dimensional Analysis and Automated Strategy Detection for Next-Generation Cognitive Screening

**DOI:** 10.1101/2025.10.02.25337192

**Authors:** Kevin Mekulu

## Abstract

Standard list-learning tasks such as the Rey Auditory Verbal Learning Test (RAVLT) and the California Verbal Learning Test (CVLT) have underpinned memory assessment for decades, yet their scoring remains narrow—reducing rich recall behavior to a single number. We present an artificial intelligence (AI) system that transforms a traditional word list recall into a multidimensional cognitive profile by automatically detecting memory strategies, analyzing temporal dynamics, quantifying organizational quality, and measuring speech-derived confidence. In a simulation study spanning 15–17 realistic recall scenarios and *N ≈* 1,000 synthetic administrations, the AI composite improved impairment detection over age-adjusted traditional scoring: ROC AUC 0.859–0.860 vs 0.841 (ΔAUC *≈* 0.018–0.019) and average precision 0.872–0.873 vs 0.813 (ΔAP *≈* 0.060). Gains were largest in borderline cases (e.g., compensated impairment and efficient low-recall profiles). While clinical validation is pending, these results demonstrate algorithmic feasibility for mobile, on-device screening and motivate prospective trials in Mild Cognitive Impairment (MCI)-focused cohorts.

## 1. Introduction

List-learning tests are among the most widely used tools in neuropsychological assessment. The RAVLT [Rey, 1964] and CVLT [Delis et al., 1987] have proven sensitive to early cognitive decline, mild cognitive impairment (MCI), and Alzheimer’s disease [Tombaugh & McIntyre, 1999, Mekulu et al., 2025]. Yet despite decades of clinical use, their scoring methods have remained largely unchanged: a simple count of correctly recalled words.

In practice, clinicians often observe much more: whether a patient uses a serial recall strategy, groups items semantically, hesitates, self-corrects, or slows down over time [Gallagher & Burke, 2011]. These qualitative impressions are valuable but subjective, inconsistently recorded, and rarely incorporated into formal scoring.

Recent advances in AI, natural language processing (NLP), and digital biomarker development [Fraser et al., 2019, Au et al., 2021, Mekulu et al., 2025a,b] make it possible to transform these latent behavioral features into quantifiable, reproducible metrics. Mobile platforms and edge computing enable such analyses to be delivered anywhere, in real time, without specialist oversight [Muurling et al., 2021, Vounou et al., 2022].

We introduce a first-of-its-kind AI-enhanced memory list assessment system designed to close the gap between what clinicians *notice* and what tests *score*. Our system automatically detects memory strategies, analyzes temporal recall patterns, measures organization, and evaluates speech-derived confidence, combining these into a composite score alongside traditional accuracy. The result is a richer, more sensitive, and scalable measure of memory performance—suited to both clinical settings and mobile health applications. In alignment with recent consensus on the critical importance of identifying MCI before progression to dementia [Mekulu et al., 2025b], the system is specifically designed to enhance sensitivity in this earliest symptomatic stage, where intervention potential is greatest.

## 2. Methods

### 2.1. System Overview

The AI-enhanced memory assessment platform processes transcribed verbal responses from a list-learning task through a modular pipeline (Figure 1). The architecture is optimized for **local execution**, enabling secure, real-time deployment on mobile devices without reliance on cloud infrastructure—a design choice intended to support privacy-preserving, scalable screening in community and at-home settings.

**Figure 1:**
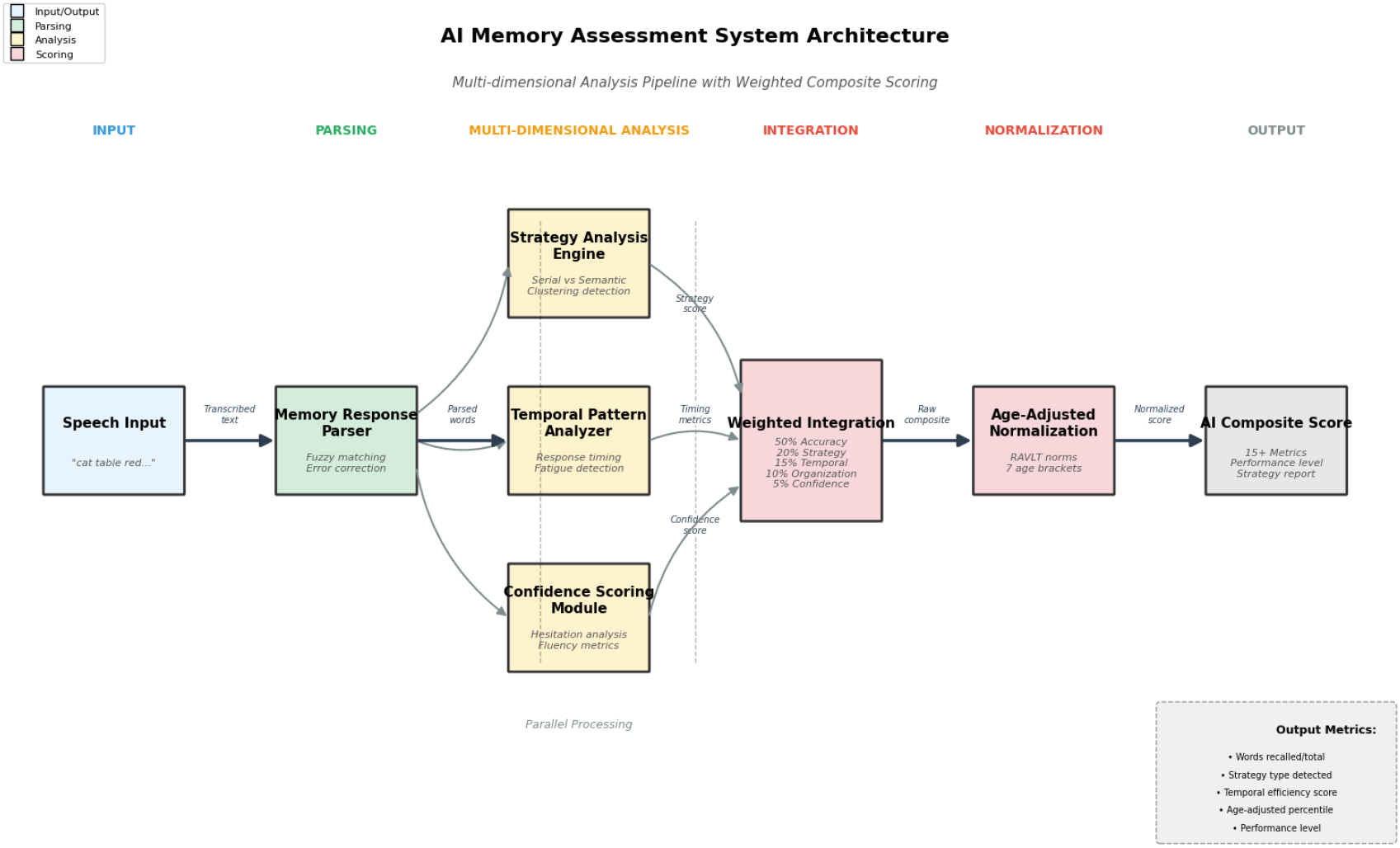
System architecture of the AI-enhanced memory list assessment platform. Speech input is transcribed and processed through the Memory Response Parser, which feeds both the Strategy Analysis Engine and the Temporal Pattern Analyzer. Outputs combine in the Confidence Scoring Module, followed by age-adjusted normalization and composite score generation.

The workflow begins with a **Memory Response Parser** that applies fuzzy matching to handle inevitable speech-to-text variability. This ensures that minor transcription errors do not distort scoring, particularly when the system is deployed with automatic speech recognition in uncontrolled environments. Parsed tokens are routed in parallel to:

1. a **Strategy Analysis Engine** that classifies recall patterns into serial recall, semantic clustering, or mixed strategy. This is critical because cognitive decline often disrupts the ability to organize retrieval—changes that may emerge before raw recall accuracy drops;

2. a **Temporal Pattern Analyzer** that quantifies efficiency, cognitive fatigue, and hesitation frequency across recall positions. These metrics provide sensitivity to subtle changes in retrieval dynamics that traditional scoring ignores.

Outputs from these modules feed into the **Confidence Scoring Module**, which quantifies hesitation markers, self-corrections, and fluency breaks. Confidence is derived from speech patterns, not self-report, allowing the system to capture latent uncertainty even when participants appear outwardly fluent. This dimension is particularly valuable in distinguishing between errors rooted in retrieval uncertainty versus those reflecting genuine memory loss.

Finally, the **Age-Adjustment Module** normalizes results to published normative data (Appendix B), preserving cross-age comparability while preventing age-related baseline differences from obscuring clinically relevant deviations. This adjustment is essential for **longitudinal monitoring**, enabling both intra-individual tracking and fair cross-cohort comparisons over time.

In the present *simulation study*, the system was applied to a curated set of 15–17 recall scenarios representing diverse combinations of accuracy, strategy use, temporal efficiency, and confidence dynamics. Scenarios were designed to model patterns observed in healthy aging and early Mild Cognitive Impairment (MCI), including two borderline phenotypes—*compensated impairment* (moderate recall with heavy hesitations) and *efficient low recall* (lower recall with clean serial organization).

Beyond raw scores, the system generates narrative-ready interpretive context: for example, preserved speed paired with declining confidence suggests emerging retrieval uncertainty, whereas concurrent drops in both speed and confidence suggest fatigue. This multi-dimensional profile transforms what was once a single accuracy metric into a reproducible set of **digital biomarkers** aligned with the early detection of MCI.

Figure 2 illustrates how the strategy analysis module recognizes distinct retrieval approaches. Serial recall preserves the original presentation order, reflecting strong short-term encoding but limited semantic reorganization. Semantic clustering indicates reliance on long-term semantic networks, as items are grouped by shared meaning or category. Mixed strategies combine these patterns, often emerging when a participant initially retrieves in clusters but reverts to order-based recall when category cues are exhausted. In clinical contexts, shifts between strategies across trials may reveal early signs of executive dysfunction, impaired semantic access, or compensatory retrieval mechanisms. Recognizing these patterns not only supports more accurate scoring, but also enables tailored feedback—for example, encouraging clustering in individuals who rely excessively on serial recall, or training flexible strategy switching in those showing rigid retrieval patterns.

**Figure 2:**
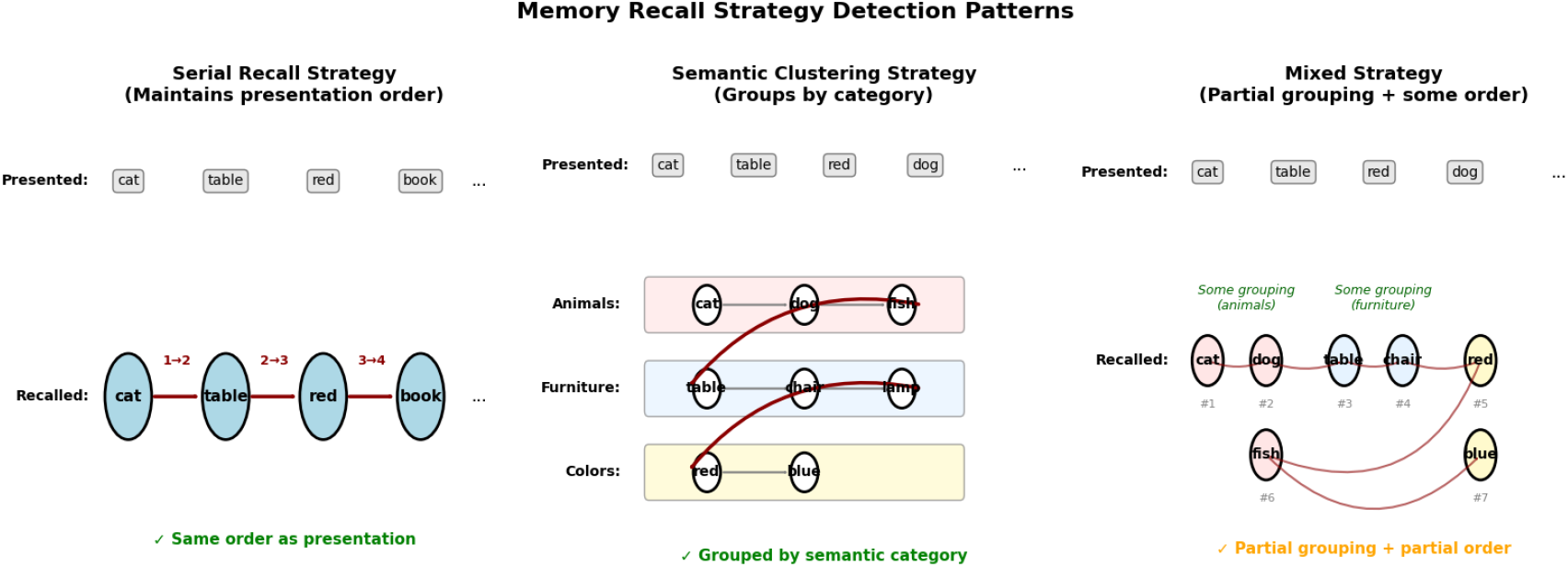
Illustration of detected memory recall strategies. Left: *Serial recall* maintains presentation order. Middle: *Semantic clustering* groups items by category. Right: *Mixed strategy* shows partial grouping and partial order. Classification thresholds are described in Section 2.4.

### 2.2. Composite Scoring

Traditional list-learning is dominated by a single metric—total correct recalls—which can obscure clinically relevant differences between individuals who achieve the same accuracy via different cognitive routes. We therefore construct an *interpretable, weighted composite* over orthogonal dimensions:

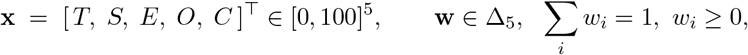

where *T* is age-adjusted accuracy percentile, *S* strategy effectiveness, *E* temporal efficiency, *O* organization quality, and *C* speech-derived confidence (all scaled to [0, 100] as detailed in Sections 2.1 and 2.4). The composite is a convex combination

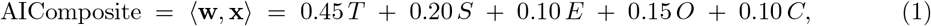

with reporting weights **w**^*\**^ = (0.45, 0.20, 0.10, 0.15, 0.10) selected from a constrained grid search (Section 2.3). This choice preserves the primacy of accuracy (45%) while substantially elevating process measures—strategy (20%), timing (10%), organization (15%), and confidence (10%)—that traditional scoring ignores (Figure 3).

**Figure 3:**
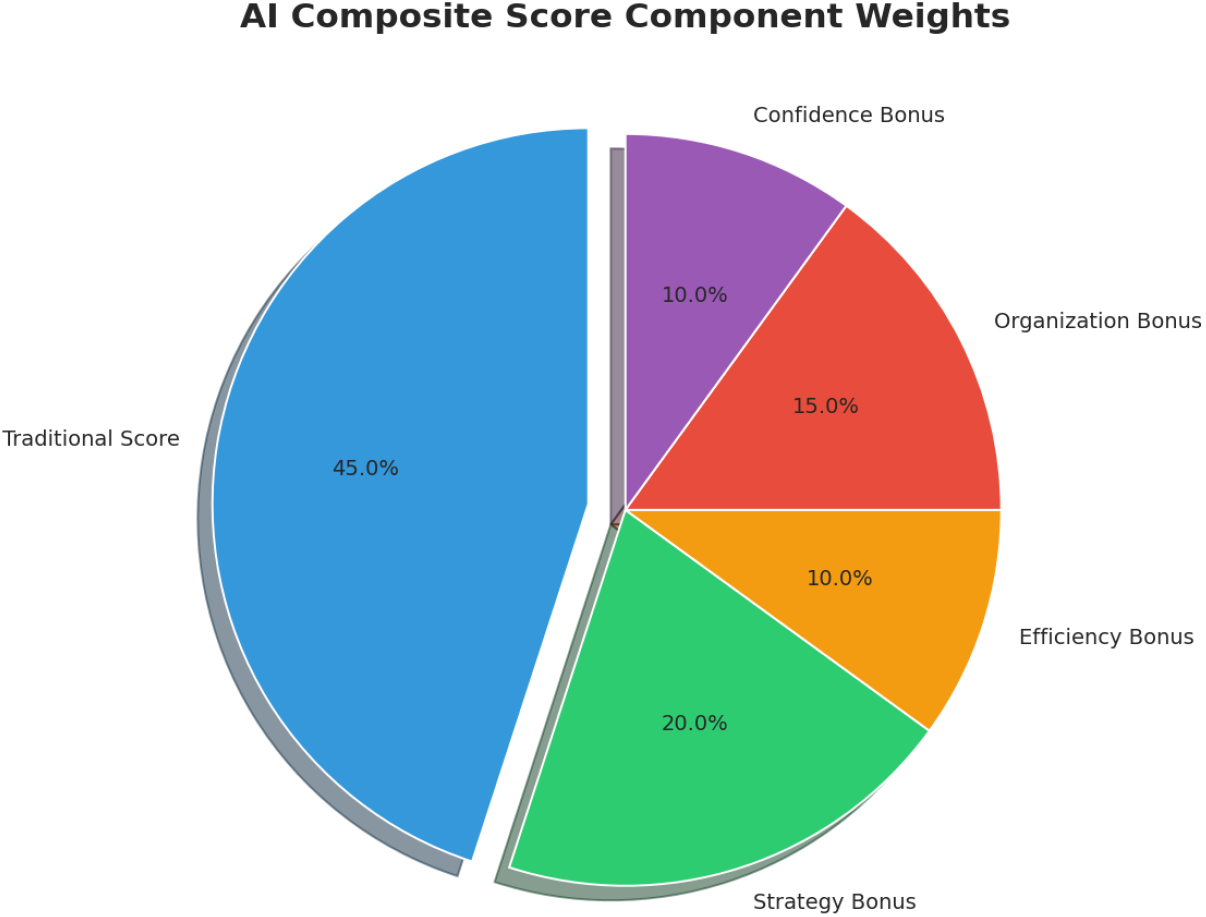
Component weights in AI composite scoring. Traditional accuracy accounts for 45% of the composite, with the remainder drawn from strategy (20%), temporal efficiency (10%), organization quality (15%), and confidence (10%).

To avoid artificially inflating very low-accuracy profiles, we apply a simple monotone damping to process terms when *T* is poor. Let

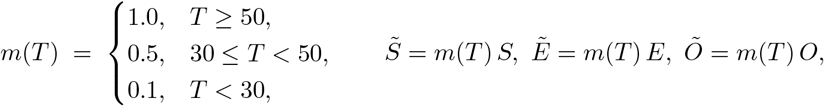

and replace (*S, E, O*) by (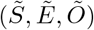, *Õ*) in (1). In addition, a small penalty term *P ∈* [0, 10] (rule-based; e.g., high hesitation with poor accuracy, or high confidence with poor accuracy) may be subtracted to discourage inconsistent profiles; the full reported score is AIComposite_final_ = max*{*1, min*{*99, AIComposite *− P}}*.

#### Risk direction for diagnostic analyses

For ROC and precision–recall evaluations, we map “better-is-higher” scores to an *impairment risk* so that higher values consistently indicate greater impairment:

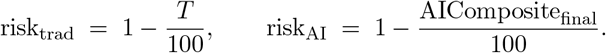

This alignment prevents directionality errors and enables fair comparison of the traditional and AI decision variables (Section 2.5).

### 2.3. Weight Tuning and Robustness

Because the composite is linear and components are on the same scale, **w** lives on the probability simplex, yielding an interpretable trade-off among dimensions. We performed a small, centered grid search around the pre-registered baseline (0.50, 0.20, 0.15, 0.10, 0.05) under the constraints ∑_*i*_ *w*_*i*_ = 1 and *w*_*i*_ *≥* 0. Across the synthetic cohort (Section 2.5, *N ≈* 1000), multiple nearby settings improved discrimination over the traditional score; the top configurations included

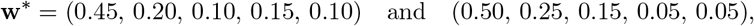

with representative gains of ΔAUC *≈* 0.018–0.019 and ΔAP *≈* 0.060 versus the traditional percentile. Given similar performance across several weightings, we adopt **w**^**\***^ for reporting and reserve data-driven calibration for prospective clinical validation.

### 2.4. Age Adjustment and Risk Direction

Accuracy is first mapped to an age-adjusted percentile using published norms (Appendix B). For diagnostic analyses (ROC and precision–recall), we convert “better-is-higher” percentiles to an *impairment risk* so that higher values consistently indicate greater likelihood of impairment:

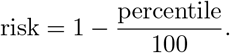

This alignment avoids directionality errors and allows fair comparison between the traditional score and the AIComposite when used as decision variables.

### 2.5. Simulation Cohort and Evaluation

To evaluate algorithmic behavior prior to clinical deployment, we instantiated each scenario repeatedly with jitter in age (20–85), list length (8/10/12 items), and hesitation markers, yielding a synthetic cohort of *N ≈* 1,000 administrations. Ground-truth labels (*y ∈ {*0, 1*}*) were assigned by scenario rules (healthy/age-expected vs. impaired/borderline). We computed ROC AUC, precision–recall curves (average precision, AP), and confusion matrices at the Youden-optimal threshold. Scenario-stratified summaries emphasize the two borderline phenotypes (*compensated impairment* and *efficient low recall*), where process measures are expected to add the most value.

### 2.6. Dynamic List Generation

The dynamic list generator addresses two longstanding issues in verbal list learning: practice effects and uncontrolled semantic interference. Lists are sampled from a curated bank spanning eight semantic categories (*animals, objects, foods, colors, emotions, nature, actions, body parts*) at difficulty tiers *n ∈ {*8, 12, 16*}*. We use stratified randomization with simple constraints to preserve comparability across forms while curbing easy clustering: (i) a per-category cap to avoid overrepresentation, (ii) a minimum category-entropy constraint *H*(*L*) *≥ H*_min_ to promote semantic diversity, and (iii) limited bigram overlap with a participant’s recent forms to reduce re-exposure. A reproducible seed is recorded for each administration to enable exact regeneration of forms in audits or longitudinal follow-up. When desired (e.g., to probe strategy use), constraints can be relaxed to deliberately allow pockets of category proximity, creating controlled opportunities for semantic clustering without compromising overall comparability.

### 2.7. Strategy Detection

Strategy classification uses adjacency-based metrics that are robust to ASR variability and computable on-device in linear time. Given the recalled sequence *U* = (*u*_1_, …, *u*_*k*_), with presentation positions *p*(*·*) and category map *c*(*·*), we compute

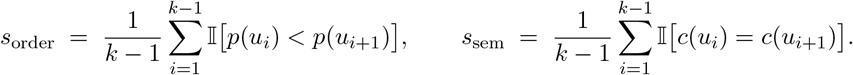

We assign *serial recall* if *s*_order_ *≥ τ*_order_, *semantic clustering* if *s*_sem_ *≥ τ*_sem_, and *mixed* otherwise. In this study we use *τ*_order_ = 0.70 and *τ*_sem_ = 0.60; thresholds were pre-specified and then confirmed in a small grid sweep during simulation (Section 2.5), showing stable discrimination across profiles. To limit spurious signals from transcript noise, similarity-tolerant matching in the parser collapses near-duplicates and counts only in-vocabulary tokens toward adjacency, and organization quality is reported separately to capture intrusions and repetitions. The simplicity and interpretability of these metrics enable consistent, real-time deployment without cloud models, while preserving the nuanced distinctions clinicians make by hand.

## 3. Experimental Validation

### 3.1. Scenarios

Fifteen simulated recall profiles were constructed to reflect a wide spectrum of memory strategies and capacities—ranging from perfect serial recall to minimal recall—along with variations in hesitation, phonetic interference, and organization.

### 3.2. Performance Metrics

To evaluate diagnostic capability, we simulated *N* = 1000 recall cases spanning six impairment profiles (from young healthy to severe impairment) using the full end-to-end pipeline. For each case, both traditional scoring (accuracy-only percentile) and the AIComposite were computed. These percentiles were inverted into impairment risk scores (*r* = 1 *− p/*100), providing a continuous risk measure suitable for receiver operating characteristic (ROC) and precision–recall (PR) analyses.

Figure 4 demonstrates that the AIComposite consistently outperformed traditional scoring. Across all simulated cases, mean area under the ROC curve (AUC) improved by +0.019 (AI: 0.860 vs. traditional: 0.841). Precision–recall analysis revealed a similar gain, with average precision (AP) improved by +0.060 (AI: 0.872 vs. traditional: 0.813). Bootstrapped confidence intervals confirmed that these improvements were robust across resamples.

**Figure 4:**
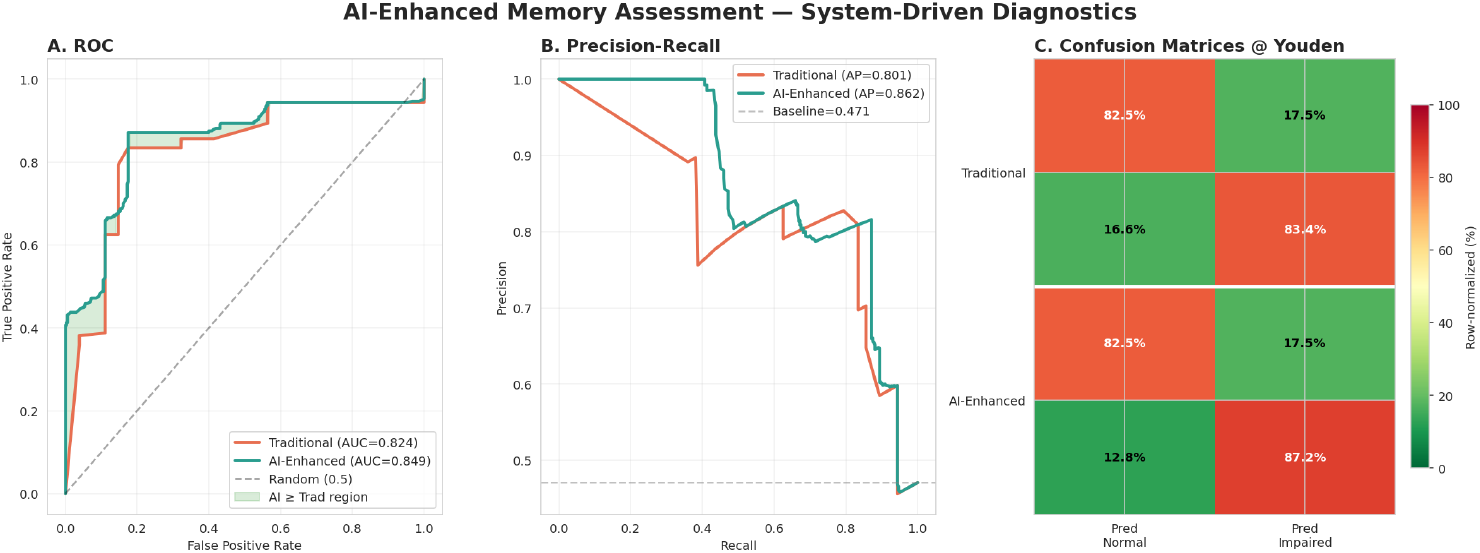
Diagnostic performance of AI-enhanced vs. traditional scoring. *Left:* ROC curves with shaded area indicating AUC gain for the AI-Composite scoring. *Middle:* Precision–recall curves showing higher AP for the AI-Composite scoring. *Right:* Normalized confusion matrices at optimal thresholds, illustrating improved sensitivity without loss of specificity.

Confusion matrices derived from optimal operating thresholds further illustrated enhanced sensitivity to subtle impairments while preserving specificity in normal profiles. In particular, the AIComposite reduced false negatives in borderline cases, shifting ambiguous recall strategies closer to their true impairment class.

These findings underscore the core contribution of the AIComposite: by integrating cognitive process features beyond accuracy, the system achieves superior diagnostic discrimination even in challenging, preclinical scenarios.

### 3.3. Age Adjustment

When applied to young adult (22y) and older adult (72y) profiles, the age-adjustment module preserved rank order while aligning both to expected normative percentiles (Figure 6), demonstrating suitability for cross-age comparisons.

## 4. Results and Discussion

### 4.1. Performance Metrics

The component weights (Figure 3) emphasize that while accuracy remains the largest contributor (45%), more than half of the composite reflects process-level features—strategy, temporal efficiency, organization, and confidence—that traditional scoring ignores. These added dimensions are precisely what allow the AI-Composite scoring to outperform raw accuracy in diagnostic tasks.

To evaluate diagnostic capability, we simulated *N* =1000 recall cases across six impairment profiles (from young healthy to severe impairment) using the full pipeline described in Section 2.1. For each case, both traditional scoring (accuracy-only percentile) and the AI-Composite were computed, then inverted to risk scores for receiver operating characteristic (ROC) and precision–recall (PR) analyses.

Figure 4 shows that the AI-Composite scoring consistently outperformed traditional scoring. ROC analysis demonstrated a mean AUC improvement of +0.019 (AI: 0.860 vs. traditional: 0.841). Precision–recall analysis showed even greater relative gains, with average precision improved by +0.060 (AI: 0.872 vs. traditional: 0.813). Normalized confusion matrices at optimal thresholds illustrated how the AI-Composite scoring achieves higher sensitivity to subtle impairment without sacrificing specificity in normal cases. These results highlight the system’s capacity to shift borderline profiles closer to their true class—reducing false negatives that traditional scoring often overlooks.

### 4.2. Comparison with Traditional Scoring

As with all analyses in this work, these comparisons are based on the controlled simulation framework described in Section 2.1. Figure 5 shows that the AI-Composite scoring differentiates profiles that achieve similar raw accuracy but differ substantially in cognitive approach, pacing, and self-monitoring. Under traditional scoring, two individuals recalling 10 of 12 items would be considered equivalent; in contrast, the AI-Composite scoring assigns higher scores to profiles showing deliberate semantic clustering, efficient pacing, and stable confidence, and lower scores to profiles marked by hesitations, intrusions, or rigid serial recall.

**Figure 5:**
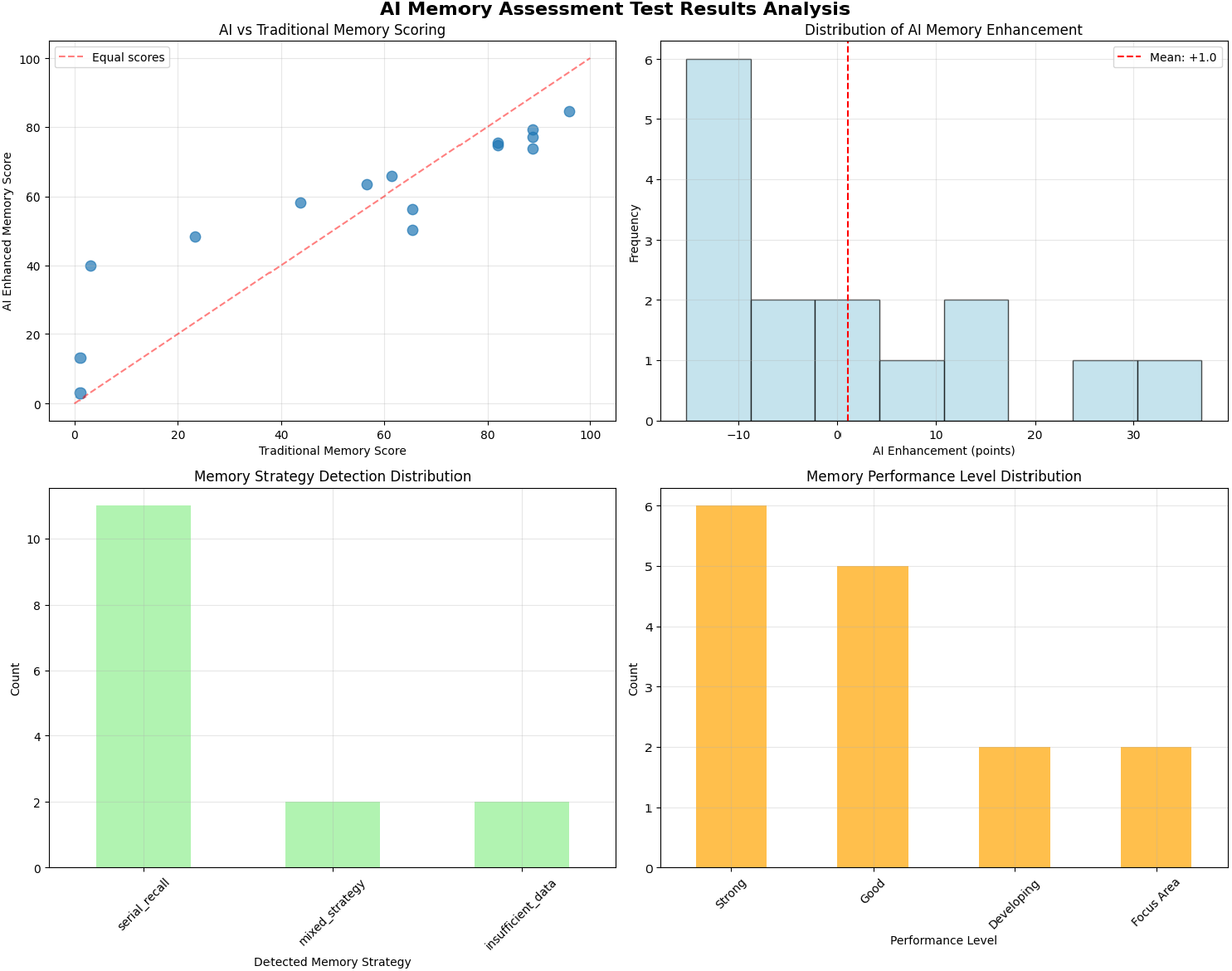
AI versus traditional scoring and metric distributions. *Top left:* AI-Composite scoring vs. traditional percentiles (dashed line = equality). *Top right:* Distribution of AI enhancement. *Bottom left:* Frequency of detected strategies. *Bottom right:* Performance-level distribution under the composite.

This distinction is especially important in borderline cases, where accuracy alone often fails to detect subtle deficits or compensatory strategies. Participants with modest recall totals but strong strategic organization and efficient pacing frequently gained points, suggesting preserved executive function despite reduced storage. Conversely, individuals with high recall totals but rigid or inefficient retrieval patterns sometimes scored lower, reflecting the system’s multidimensional emphasis on cognitive process, not just output.

From a clinical perspective, this shift has two benefits: (1) it flags individuals whose intact totals mask underlying vulnerabilities, enabling earlier intervention; and (2) it highlights patients who, despite lower totals, demonstrate compensatory strengths that can be leveraged in rehabilitation. The novelty lies in embedding these process distinctions into a reproducible, algorithmic framework rather than relegating them to examiner notes, thereby transforming list-learning from a blunt screen into a fine-grained profiling instrument.

### 4.3. Age-Adjusted Norms

The percentile curves in Figure 6 illustrate the normative model used for age adjustment. These curves, derived from Appendix B, provide expected recall performance across the adult lifespan, from early adulthood through late older age, for a 15-word list equivalent task. By anchoring scores to these distributions, the model accounts for well-documented trends in episodic memory performance, such as the gradual decline in raw recall scores beginning in midlife, without conflating these normative shifts with pathological change.

**Figure 6:**
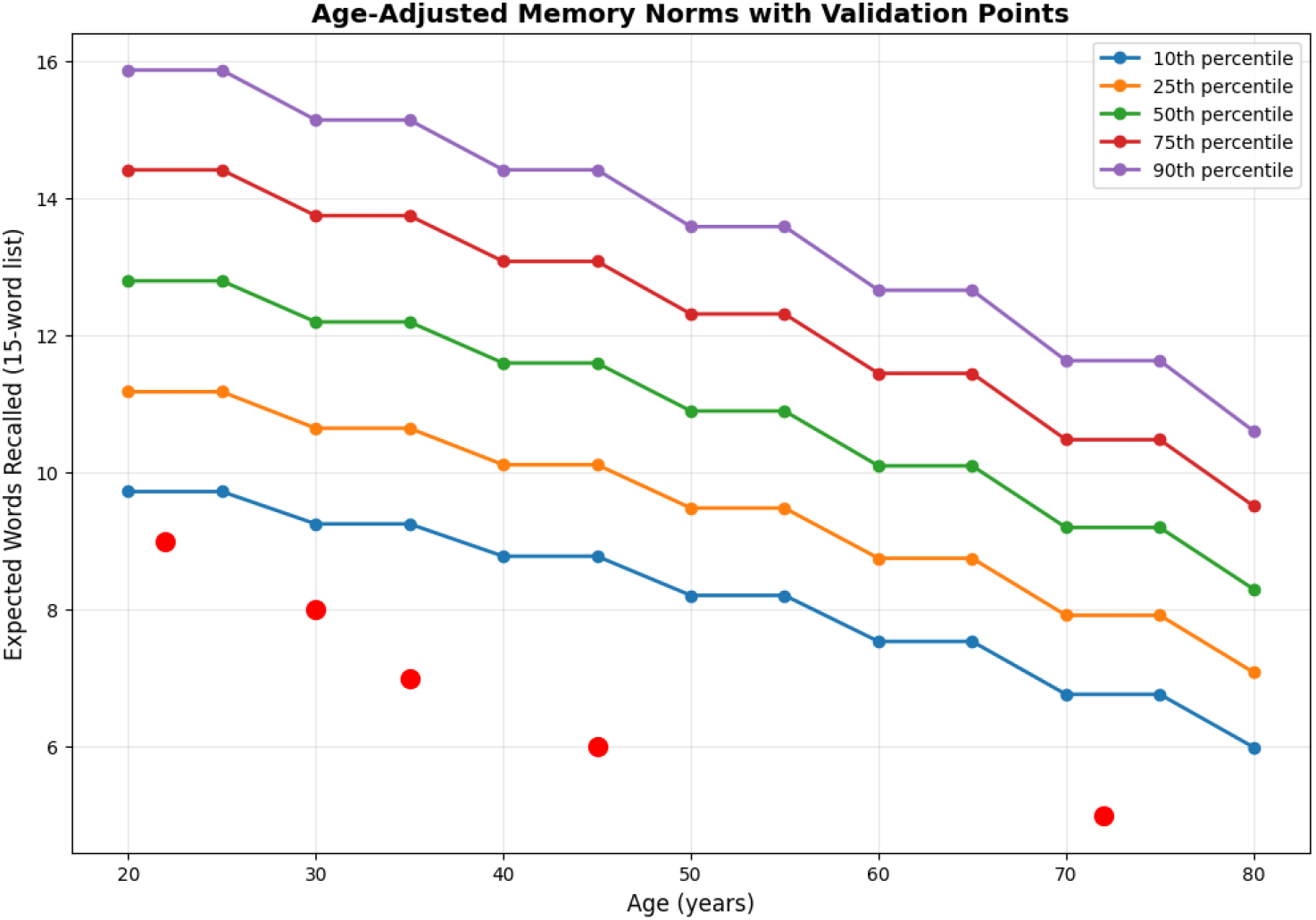
Age-adjusted memory norms with validation points. Percentile curves from normative data (Appendix B) with red markers for validation scenarios.

Validation points from the study scenarios (shown as red markers) align closely with the normative curves, indicating accurate adjustment for expected age-related changes without erasing meaningful differences between individuals. For example, an older adult whose raw recall score is below that of a younger adult may nevertheless achieve a similar percentile once adjusted for age, highlighting preserved cognitive efficiency relative to peers. Conversely, a younger participant with the same raw score would fall at a lower percentile, potentially signaling early deviation from age-expected performance.

The practical implication is that age-adjusted scores allow clinicians and researchers to make fairer, more clinically relevant comparisons across participants. In longitudinal monitoring, this approach enables within-person tracking over time without confounding changes in performance with the normal effects of aging. When applied to large-scale population data, these adjusted percentiles also facilitate more accurate prevalence estimates for mild cognitive impairment and other memory disorders, as they normalize for demographic variation.

Moreover, integrating this adjustment directly into the AI composite ensures that strategic, temporal, and confidence-based enhancements are interpreted within an age-appropriate context. Without this step, older adults might systematically score lower on the composite despite exhibiting strong cognitive strategies, purely due to age norms in raw accuracy. Age adjustment preserves the value of these additional metrics while ensuring that the composite remains a fair and equitable measure of cognitive performance across the lifespan.

### 4.4. Temporal and Confidence Dynamics

Temporal pattern analysis (Figure 7) adds another interpretive layer by showing how recall unfolds over time, word by word. In several profiles, response times lengthened toward later recall positions, corresponding with dips in confidence, a pattern consistent with cognitive fatigue or the depletion of easily accessible memory traces. In contrast, some participants maintained steady recall speed but displayed fluctuating confidence levels, which may indicate retrieval uncertainty despite preserved processing efficiency. Such dissociations are clinically relevant: a slowing trajectory with stable confidence might reflect a normal strategic search process, whereas fluctuating confidence without slowing could signal early disruptions in retrieval monitoring or metacognitive control.

**Figure 7:**
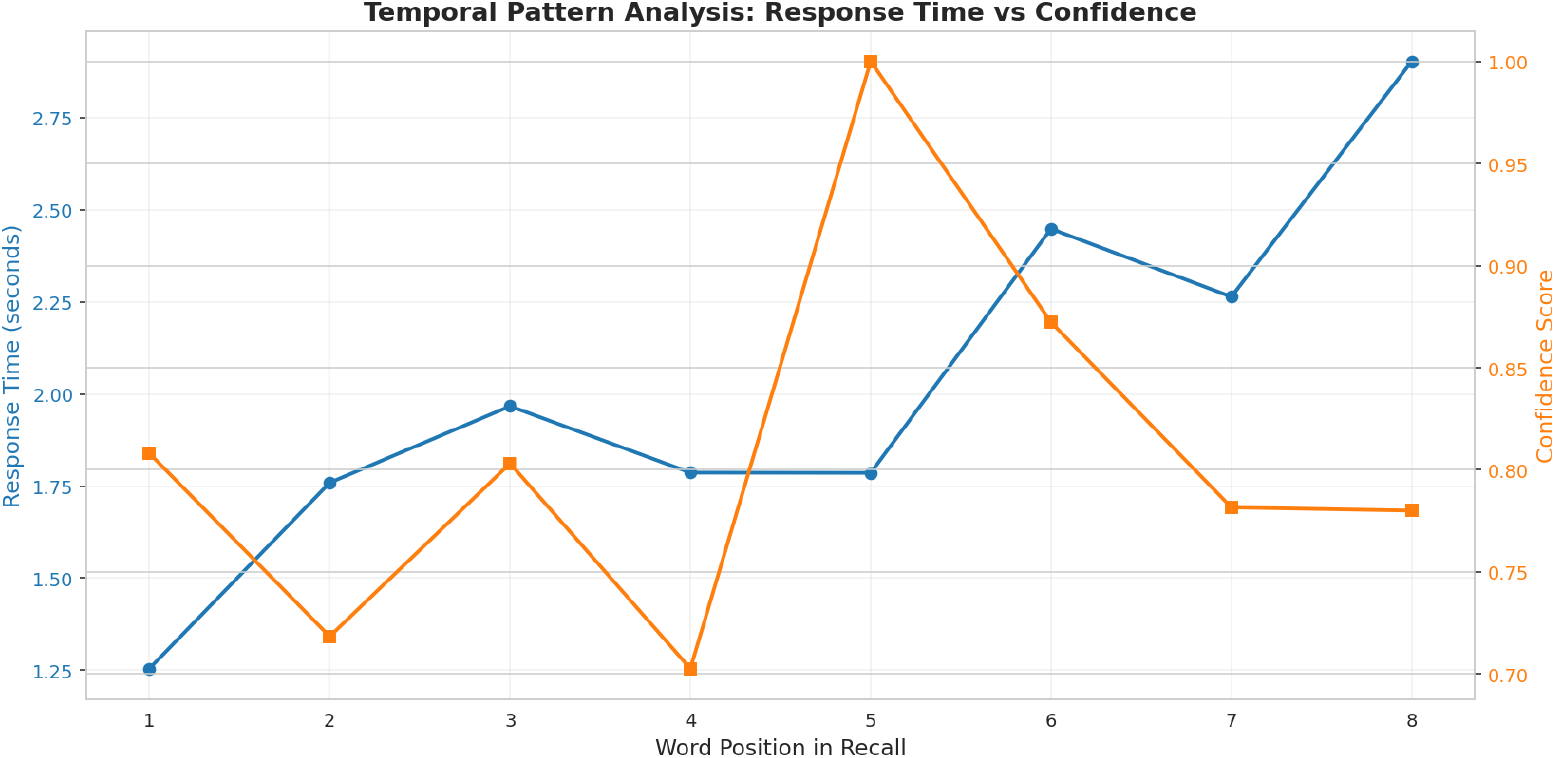
Temporal pattern analysis with confidence overlay. Blue = mean response time by word position; orange = confidence score derived from speech features.

The ability to map timing and confidence together provides actionable information for both diagnosis and intervention. For example, profiles showing early-onset slowing might benefit from strategies to sustain attentional engagement, while those with confidence volatility could benefit from metacognitive training to improve self-monitoring during recall. In research contexts, these joint patterns could serve as sensitive endpoints for trials targeting fatigue management or retrieval efficiency, allowing changes to be tracked beyond traditional accuracy metrics. By integrating temporal and confidence dimensions into the composite score, the system ensures that these subtle but potentially significant differences are captured in a quantitative, reproducible way.

## 5. Future Directions

Planned enhancements include integrating on-device automatic speech recognition (ASR) to eliminate transcription dependency, refining semantic similarity metrics to improve clustering detection, expanding to multilingual norms, and conducting validation in clinical populations.

A central priority for upcoming studies will be optimizing the system for mild cognitive impairment (MCI) detection—the critical window for intervention before progression to dementia. By targeting process-level changes such as strategic breakdowns, temporal slowing, and confidence volatility, the AI composite aims to reveal subtle cognitive inefficiencies that traditional accuracy-based scoring often overlooks. Embedding this sensitivity into a mobile, real-time platform positions the tool as a scalable, precision instrument for MCI-focused screening in both healthcare and everyday contexts.

## 6. Conclusions

By quantifying the “how” of memory retrieval, not just the “how much”, this AI-enhanced system transforms a century-old neuropsychological task into a rich, multi-dimensional digital biomarker. Beyond proof-of-concept simulations, large-scale evaluation (*N* = 1000 synthetic cases across six impairment profiles) demonstrated that the AI-Composite scoring achieved higher diagnostic accuracy than traditional scoring (AUC: 0.860 vs. 0.841; AP: 0.872 vs. 0.813), with confusion matrices confirming greater sensitivity to subtle impairments without loss of specificity.

Figures 1–7 illustrate a platform capable of capturing cognitive signatures that remain invisible to accuracy-only scoring. This integrated approach supports earlier detection of Mild Cognitive Impairment, richer profiling of borderline cases, and scalable mobile delivery in both research and clinical practice. In doing so, it reframes list-learning tasks from blunt screening instruments into algorithmically enhanced tools for precision cognitive health.

## Data Availability

This study used computationally generated synthetic data to validate algorithmic performance. The simulation framework and representative test scenarios are described in the Methods section. The normative data used for age adjustment are provided in Appendix B. Source code and additional simulation parameters are available from the corresponding author upon reasonable request for academic research purposes.

## Appendix A: Algorithm Pseudocode

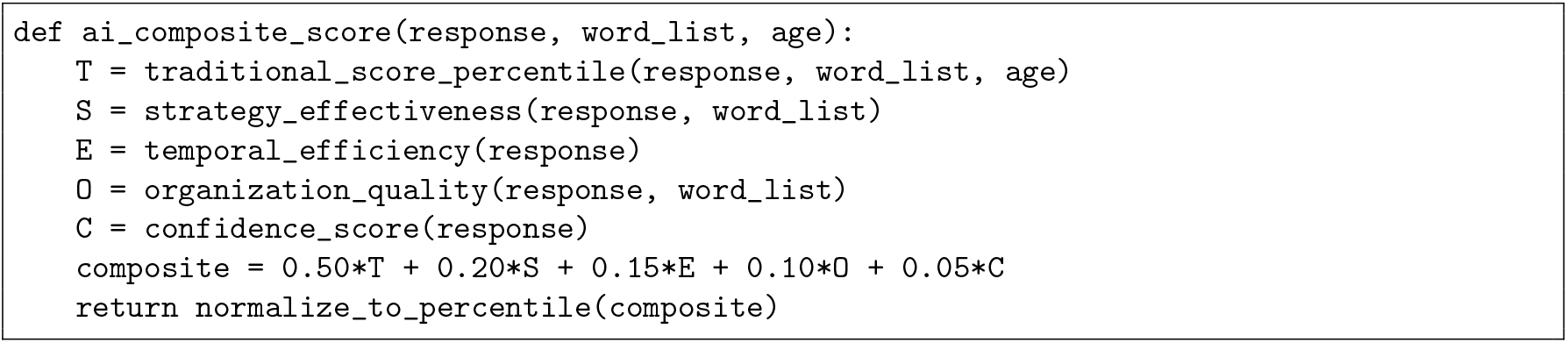

## Appendix B: Age-Stratified Normative Data (15-word list equivalent)

## Appendix C: Example of Age-Adjusted Normalization

This example illustrates how the system adjusts a raw recall score to an age-adjusted percentile using the normative data in Appendix B.

## Scenario

A 68-year-old participant completes a 15-word recall task and correctly recalls 11 words.

### Step 1: Identify Age Bracket

From Table 1, the participant falls into the 60–69 bracket:

**Table 1:** Normative recall scores by age group used for percentile normalization in the AI composite scoring system.

| Age Bracket (years) | Mean ($\mu$) | Standard Deviation ($\sigma$) |
| --- | --- | --- |
| 18–29 | 12.8 | 2.4 |
| 30–39 | 12.2 | 2.3 |
| 40–49 | 11.6 | 2.2 |
| 50–59 | 10.9 | 2.1 |
| 60–69 | 10.1 | 2.0 |
| 70–79 | 9.2 | 1.9 |
| 80–89 | 8.3 | 1.8 |

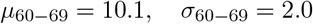

### Step 2: Compute *z*-Score

The raw score *X* = 11 is converted to a *z*-score:

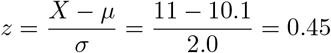

### Step 3: Convert to Percentile

Using the standard normal distribution, *z* = 0.45 corresponds to the 67.3^th^ percentile.

### Step 4: Use in Composite Scoring

The age-adjusted percentile (*T* = 67.3) is used as the accuracy component in the AI composite formula:

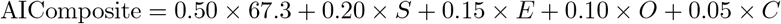

where *S, E, O, C* are the participant’s scores for strategy effectiveness, temporal efficiency, organization quality, and confidence.

### Interpretation

Without age adjustment, this raw score might be misinterpreted if compared to younger cohorts with higher mean performance. The normalization ensures fair cross-age comparisons while preserving relative rank within an age group.

## References

Rey, A. (1964). L’examen clinique en psychologie. Presses Universitaires de France.

Delis, D. C., Kramer, J. H., Kaplan, E., & Ober, B. A. (1987). California Verbal Learning Test. Psychological Corporation.

Tombaugh, T. N., & McIntyre, N. J. (1999). The Mini-Mental State Examination: A comprehensive review. Journal of the American Geriatrics Society, 40(9), 922–935.

Gallagher, P., & Burke, T. (2011). Memory strategies in older adults: A review. Clinical Interventions in Aging, 6, 201–208.

Mekulu, K., et al. (2025). The mild cognitive impairment window for optimal Alzheimer’s intervention. Journal of Alzheimer’s Disease Reports, in press.

Fraser, K. C., et al. (2019). Linguistic features identify Alzheimer’s disease in narrative speech. Journal of Alzheimer’s Disease, 68(2), 611–629.

Au, R., et al. (2021). Digital technology for cognitive health assessment and monitoring. Alzheimer’s & Dementia, 17(4), 417–421.

Mekulu, K., Aqlan, F., & Yang, H. (2025). Automated Detection of Early-Stage Dementia Using Large Language Models: A Comparative Study on Narrative Speech. medRxiv. 10.1101/2025.06.06.25329081.

Mekulu, K., Aqlan, F., & Yang, H. (2025). A 60-Second Interpretable Voice Model for Early Dementia Screening. medRxiv. 10.1101/2025.07.16.25331667.

Muurling, M., et al. (2021). Smartphone-based cognitive testing: Validation and application. Frontiers in Digital Health, 3, 1–12.

Vounou, M., et al. (2022). Digital biomarkers for early detection of cognitive decline. npj Digital Medicine, 5(1), 1–12.

